# Bivariate Multiscale Sample Entropy of Frontoparietal–Subgenual ACC Dynamics in Depression During Accelerated TMS at 7 T

**DOI:** 10.64898/2026.11.12.26367710

**Authors:** Niklaus Denier, Matthias Grieder, Kay Jann, Pascal Borel, Stefan Dendorfer, Gianluca Florineth, Kristina Adorjan, Andrea Federspiel, Roland Wiest, Leila M. Soravia, Sebastian Walther, Tobias Bracht

## Abstract

**Objective:** To examine whether longitudinal changes in frontoparietal (FP)–subgenual anterior cingulate cortex (sgACC) joint temporal complexity differ between sgACC hemispheres in patients undergoing accelerated TMS (aTMS) and untreated healthy controls.

**Methods:** Twenty-four patients with current depressive episodes and fifteen healthy controls underwent 7T resting-state fMRI at baseline and follow-up. Patients received aTMS localized to the left DLPFC using Beam-F3. The same bilateral FP time series was paired separately with left and right sgACC time series. Bivariate multiscale sample entropy (bMSE) across scale factors a1–a10 was summarized by trapezoidal integration as a complexity index (bMSE-CI). The primary test was the Group × Time × sgACC Hemisphere interaction in a mixed repeated- measures ANOVA. Scale-specific interactions and clinical Spearman correlations were evaluated with Benjamini–Hochberg false discovery rate correction. Change scores were follow-up minus baseline.

**Results:** The bMSE-CI Group × Time × Hemisphere interaction was significant (F(1,37) = 6.043, p = 0.0188, partial η^2^ = 0.140). Mean left/right bMSE-CI changes were −0.286/+0.124 in patients and +0.323/+0.112 in controls. Of ten scale-specific three-way interactions, only a2 survived FDR correction (q = 0.0435). No baseline or change-score bMSE–MADRS association survived the specified FDR corrections. A random-intercept-only mixed-model sensitivity analysis did not reach statistical significance for the three-way interaction (p = 0.0718).

**Conclusion:** FP–sgACC joint temporal complexity showed hemisphere-dependent longitudinal differences between patients undergoing aTMS and healthy controls. These exploratory findings motivate further evaluation of bMSE as a candidate measure of longitudinal neural change in depression. Independent replication, more precise reliability estimates, and prospective validation of clinical relevance are needed.

## Introduction

Repetitive transcranial magnetic stimulation (rTMS) is a promising non-invasive brain stimulation method with high levels of evidence for treating major depressive disorder and bipolar depression, particularly when standard medications are ineffective (Brunoni et al., 2017; Lam et al., 2024). rTMS is usually applied to the left dorsolateral prefrontal cortex (DLPFC) and has shown strong antidepressant effects and good safety (Brunoni et al., 2017; Trapp et al., 2025). A particularly time-efficient protocol is the intermittent theta-burst stimulation (iTBS), which is equally effective as conventional rTMS protocols (Blumberger et al., 2018). Accelerated TMS protocols (aTMS) are applied several times a day and show potential to achieve treatment response in much shorter treatment periods (Caulfield et al., 2022; Cole et al., 2020; Lan et al., 2023; Ramos et al., 2025; Sheline et al., 2024; van Rooij et al., 2024). Recent randomized sham-controlled trials have demonstrated antidepressant efficacy of accelerated theta-burst stimulation in treatment-resistant unipolar and bipolar depression (Ramos et al., 2025; Sheline et al., 2024).

Structural and functional MRI studies indicate that rTMS produces significant changes in brain network connectivity, especially in mood-related regions like the DLPFC and anterior cingulate cortex (ACC) (M. D. Fox et al., 2012; Sheline et al., 2024). The subgenual ACC (sgACC) is a key region in the neurobiology of depression and an indirect target for rTMS treatments (X. Chen et al., 2025; Raij et al., 2023). Neuroimaging studies demonstrated that the sgACC acts as a central hub in emotion regulation networks and displays structural changes, such as reduced gray matter volume, along with abnormal functional connectivity (Drevets et al., 2008; Scharnowski et al., 2020). The DLPFC is a node of the frontoparietal (FP) network, which participates in executive control and interacts with affective regions such as the sgACC (Benschop et al., 2022). Network models of depression emphasize interactions among executive, default-mode and salience systems (Bertocci et al., 2023; Brakowski et al., 2017; Menon, 2011; Mulders et al., 2015; Wang et al., 2022).

Complexity analyses provide an emerging method for studying functional MRI (fMRI) data beyond traditional linear metrics, quantifying the richness and variability of neural activity over time (Zhang et al., 2016). These techniques capture temporal irregularities and nonlinear patterns in brain dynamics that conventional linear measures cannot capture, offering deeper insights into neural information processing. Among these approaches, Multiscale Sample Entropy (MSE) stands out as a particularly valuable measure of neural temporal dynamics. Multiscale Sample Entropy (MSE) is a nonlinear method that measures the complexity of fMRI brain signals by computing sample entropy (SampEn) across different temporal scales (Costa et al., 2002; Richman & Moorman, 2000). Unlike single-scale approaches, MSE provides a frame-work for quantifying temporal complexity in neural signals, capturing both local and distributed temporal dynamics (Wang et al., 2018).

Recent investigations have demonstrated that intrinsic brain networks exhibit a hierarchical organization of entropy, with intrinsic networks consistently displaying higher complexity compared to extrinsic networks across various temporal scales (Trevino et al., 2024). MSE helps to characterize the integrative and segregative capacities of neural systems. Altered MSE profiles have been linked to disrupted network organization in cognitive decline and neuropsychiatric disorders (Grieder et al., 2018; Jann et al., 2025; Zhang et al., 2024). Complexity analyses such as Higuchi’s fractal dimension have also been applied in the context of depression (Denier et al., 2024). However, MSE has been primarily used in EEG data (W. Chen et al., 2025; Jaworska et al., 2018; Xu et al., 2025). Higher MSE complexity in the left frontoparietal network has been reported, suggesting a potential role of neural compensation and executive function in depressive disorders (Lin et al., 2019). These findings emphasize the relevance of MSE as a potential marker of neuroplasticity and functional compensation in depression (Grieder et al., 2025; Zhang et al., 2024).

Conventional MSE operates on univariate time series, analyzing each voxel in isolation, and thus remains limited to regional signal dynamics rather than capturing the emergent complexity of large-scale brain networks. To address this, multivariate MSE (MMSE) extends the approach to multiple time series, capturing interdependencies and joint dynamics across brain regions (Ahmed & Mandic, 2012; Ahmed & Mandic, 2011). By constructing composite state vectors that integrate information across several time series, MMSE reflects both temporal and spatial complexity, increasing sensitivity to subtle network-level alterations (Monika Lewandowska et al., 2023; Liu et al., 2023). To the best of our knowledge, there are currently no published studies applying MMSE to fMRI data.

This study investigated longitudinal changes in the joint temporal complexity of bilateral FP and left or right sgACC time series in patients with depression undergoing aTMS and untreated healthy controls. We used bivariate multiscale sample entropy (bMSE) to quantify joint temporal complexity across scale factors a1–a10 and summarized the profiles as bMSE complexity indices (bMSE-CIs). The same bilateral FP time series was used for both sgACC sides. The primary test was the Group × Time × sgACC Hemisphere interaction for bMSE-CI, assessing whether the between-group difference in longitudinal change differed between sgACC hemispheres. The hemispheric analysis was not prospectively prespecified; no directional hypothesis was asserted. Scale-specific interaction tests and correlations with clinical outcomes were exploratory. Functional connectivity (FC), regional multiscale sample entropy (MSE), and the bilateral sgACC mean-signal readout were examined as secondary measures. These analyses characterize longitudinal FP–sgACC dynamics without establishing treatment-specific causality, directed coupling, or clinical biomarker validity.

## Methods

### Participants

All 39 participants in this observational study were enrolled at the University Hospital of Psychiatry and Psychotherapy Bern, encompassing both inpatient and outpatient populations. Eligible patients were aged 18-65 years, presented with a current depressive episode, and were scheduled for TMS therapy. The patient cohort (n = 24) included individuals with major depressive disorder and bipolar disorder with current depression (moderate to severe) according to DSM-5 criteria by the American Psychiatric Association (APA, 2013). Patients with unipolar and bipolar depression were analyzed together because they shared a current depressive episode and the study focused on longitudinal FP–sgACC network dynamics during treatment. Exclusion criteria comprised psychotic or neurological disorders, substance use disorders (excluding nicotine), claustrophobia, pregnancy, and contraindications to 7-Tesla MRI. Healthy control participants (n = 15) were additionally required to have no current or lifetime psychiatric diagnosis. All study participants provided written informed consent, and the study protocol was approved by the cantonal ethics committee of Bern, Switzerland (KEK: 2023-00134). The present analysis uses data from the same observational cohort as the sgACC seed-based functional connectivity study by Bracht et al. (2026). The analysis samples differ: the present bMSE analysis includes 24 patients and 15 healthy controls, whereas the published connectivity analysis included 22 patients and 14 controls after motion-related exclusions.

### Assessments

For diagnostic assessment of mental disorders according to DSM-5, we used the Mini International Neuropsychiatric Interview (Sheehan et al., 1998). Handedness was measured using the Edinburgh Handedness Inventory (Oldfield, 1971). Depression severity was assessed using the Montgomery Asberg Depression Rating Scale (MADRS) as an objective, clinician-administered measure (Montgomery & Asberg, 1979) and the Beck Depression Inventory-II (BDI-II) as a subjective, self-reported measure (Beck et al., 1996). In healthy controls, questionnaires were administered once on the day of the first MRI scan, whereas in depressed patients, questionnaires were administered both before and after aTMS treatment on the day of the MRI scan. Change scores (ΔMADRS, ΔbMSE, and ΔFC) were calculated as follow-up minus baseline; negative MADRS change indicates symptom improvement.

Change-score definition: *Δq* = *q _follow_*__*up*_ − *q_baseline_*. Here q denotes MADRS, bMSE, or FC.

### Accelerated TMS (aTMS)

Stimulation was delivered via a figure-8 coil using a MagPro R30 stimulator (MagVenture, Denmark). The target site (left DLPFC) was localized using the Beam-F3 method (Mir-Moghtadaei et al., 2015). Individual connectivity-guided targeting was not used. Resting motor thresh-old (RMT) was determined over the right abductor pollicis brevis muscle; stimulation intensity was set to 120% RMT (Blumberger et al., 2018).

Treatment comprised 10 treatment days (Monday–Friday over 2 weeks), with four iTBS runs per day separated by reported 15-minute inter-run intervals, yielding 40 runs in total. Each iTBS run followed the standard protocol: 50 Hz triplet bursts repeated at 5 Hz (200 ms inter-burst interval), delivered in 2-second trains repeated every 10 seconds (8-second pauses between trains), totaling 600 pulses per run (duration: 3 min 9 s). With 600 pulses per run, this yielded 2,400 pulses per treatment day and 24,000 pulses across the full treatment course.

### MRI acquisition and preprocessing

MRI data were acquired on a Siemens Magnetom Terra 7T scanner (Siemens Healthcare, Er-langen, Germany) equipped with a 32-channel head/neck coil (Nova Medical, Wilmington, MA) at the Translational Imaging Center, University Hospital Bern, Switzerland. High-resolution T1-weighted structural images were obtained using a 3D MPRAGE sequence (voxel size = 0.6 × 0.6 × 0.6 mm^3^; field of view = 240 × 240 mm^2^; matrix = 384 × 384; TR = 2500 ms; TE = 3.12 ms; TI = 1050 ms; flip angle = 7°; acquisition time = 7:40). Resting-state fMRI data were collected using a multiband echo-planar imaging (EPI) sequence (multiband acceleration factor = 4; in-plane acceleration factor = 3; 68 slices; voxel size = 1.4 × 1.4 × 1.4 mm^3^; field of view = 224 × 224 mm^2^; acquisition matrix = 160 × 160; TR = 1000 ms; TE = 24 ms). The original inspected resting-state DICOM/CSA header confirmed these sequence parameters. The analyzed functional images contained 500 volumes in 77 scans and 399 volumes in one scan, corresponding to sampled durations of 500 s and 399 s at TR = 1 s.

Resting-state fMRI data were preprocessed using CONN 22a (RRID:SCR_009550) (Whitfield-Gabrieli & Nieto-Castanon, 2012). Preprocessing steps included motion realignment with unwarping, coregistration to anatomical images, normalization to the MNI template, and spatial smoothing with a Gaussian kernel of 3 mm (FWHMkernel ≈ 2 × voxel size). The CONN processing log documents realignment and unwarping to correct motion-related susceptibility-distortion interactions. Additional field-map-based or reverse-phase-encoding distortion correction and additional 7T-specific quality-control procedures were not documented in the available processing records. Denoising used the anatomical CompCor strategy with five white-matter and five CSF components (Behzadi et al., 2007), six rigid-body motion parameters and their first temporal derivatives, and session-specific ART outlier regressors. ART thresholds were 0.9 mm for motion and 5 SD for global signal changes; flagged volumes were modeled using nuisance regressors. Linear detrending was applied, with nuisance regression followed by band-pass filtering at 0.008–0.09 Hz (RegBP); temporal despiking was disabled. Slice-timing correction was not documented in the available processing records. The retained analysis cohort contained 39 participants and 78 scans. An original participant-or scan-level motion-exclusion rule and the number excluded before formation of this cohort could not be reconstructed from the available records. Mean framewise displacement (FD) was calculated to quantify head motion over the scanning period (Power et al., 2012).

### Refined-composite bivariate multiscale entropy (bMSE)

Refined-composite bivariate multiscale entropy was computed in Python using EntropyHub cMvMSEn with Refined=True and MvSampEn, embedding dimensions m = [2, 2], unit delays, tolerance r = 0.6, and Norm=True (Ahmed & Mandic, 2011; Ahmed & Mandic, 2012). For each scan, the same denoised bilateral FP time series was paired with the left, right, or secondary bilateral sgACC time series. At each scale, the refined-composite estimator pools matching probabilities across shifted coarse-grained sequences before taking their negative natural-log ratio. MvSampEn normalizes each coarse-grained channel to unit standard deviation and evaluates composite multivariate embeddings and their expanded subspaces. Thus bMSE quantifies joint temporal complexity rather than two independent univariate entropy estimates; it does not isolate directed coupling or information exchange. Scale factors a1–a10 describe temporal aggregation of the band-pass-filtered BOLD signals (0.008–0.09 Hz). At TR = 1 s, a2 and a3 represent 2-s and 3-s aggregation windows. Inverse window duration does not define an isolated physiological frequency band, and fine-scale entropy does not establish high-frequency neural activity, criticality, or excitation–inhibition balance. Figure 2 summarizes the analysis pipeline. Coarse-graining at scale a: 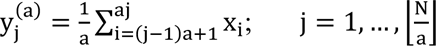. For refined-composite bMSE, the same averaging is repeated for each starting offset k = 1, …, a; each resulting channel is normalized to unit standard deviation before multivariate matching.

Two-channel embedding (unit delay): *Y*_m_^(a, k)^(*j*) = [Ỹ_1_, *j*^(a, k)^, Ỹ_1_, *j* + 1^(a, k)^, Ỹ_2_, *j*^(a, k)^, Ỹ_2_, *j* + 1^(a, k)]^. Channel 1 is FP and channel 2 is the respective sgACC signal; tildes denote normalized coarse-grained samples and m = [2, 2].

Refined-composite bivariate entropy: *bMSE*(*a*) = −*ln*[*P̅*_m+h_(*a*) / *P̅*_m_(*a*)]. P̅ denotes the matching probabilities pooled across shifted coarse-grained sequences; the expanded probability combines the channel-specific expanded subspaces as implemented by MvSampEn. The normalized-channel matching tolerance is r = 0.6.

The bilateral FP mask (FP.nii) was derived from the CONN network atlas. Left and right sgACC masks (sgACC_l.nii and sgACC_r.nii) were derived from the Automated Anatomical Labeling atlas, version 3 (AAL3). The same bilateral FP mask was paired separately with each sgACC mask. Masks were aligned to the functional image grid using their active NIfTI world-coordinate transforms and nearest-neighbor resampling. The FP mask contained 5,130 voxels (41,040 mm^3^) on the 2-mm analysis grid. The supplied left sgACC mask contained 268 voxels (2,144 mm^3^), including 216 left-sided and 52 midline voxels; the supplied right sgACC mask contained 239 right-sided voxels (1,912 mm^3^). Original mask membership was retained without additional hemispheric repartition or midline exclusion. The exact disjoint union of these masks formed the bilateral sgACC mask (507 voxels; 4,056 mm^3^), used as a secondary mean-signal readout. For each scan, BOLD signals were averaged across voxels within each aligned mask and denoised using the session-specific nuisance design and filtering described above. The identical bilateral FP time series was paired with the left or right sgACC time series to compute bMSE and Pearson functional connectivity; thus Hemisphere refers to the sgACC component only. The bilateral sgACC signal was extracted directly from the union mask rather than by averaging hemisphere-specific entropy or connectivity estimates. The masks are shown in Figure 1.

**Figure 1:**
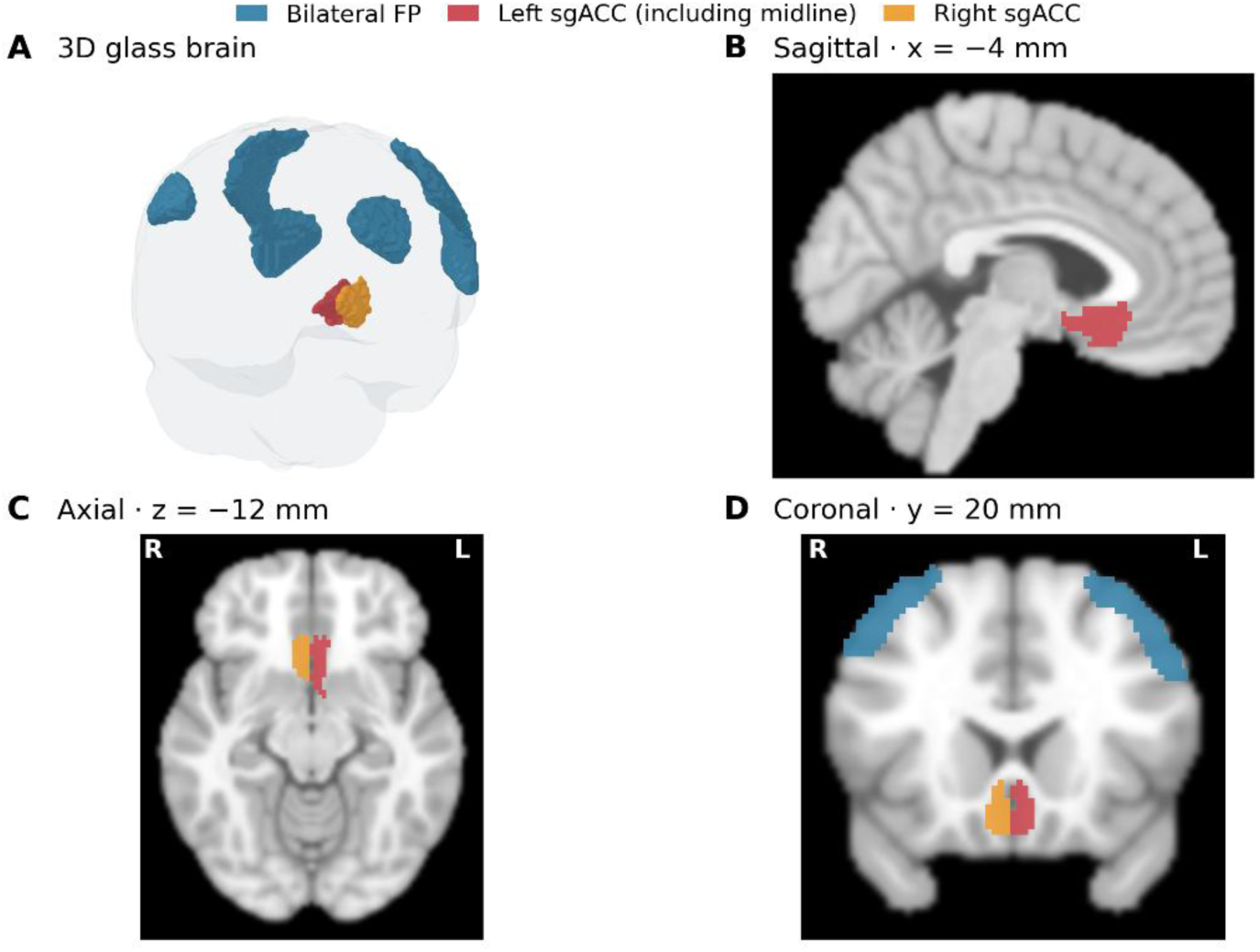
Bilateral frontoparietal network and left/right subgenual ACC regions of interest. The actual analysis masks are shown within a transparent three-dimensional MNI brain enve-lope (A; 3D glass brain) and on MNI anatomical sections: sagittal x = −4 mm (B), axial z = −12 mm (C), and coronal y = 20 mm (D). Bilateral FP is blue (5,130 voxels); supplied left sgACC is red (268 voxels, including 52 midline voxels); supplied right sgACC is orange (239 voxels). The left and right masks were retained without additional hemispheric repartition. Their exact union forms the secondary bilateral sgACC mask (507 voxels). The same FP mask is paired with each sgACC side. Section labels indicate anatomical left (L) and right (R).

**Figure 2:**
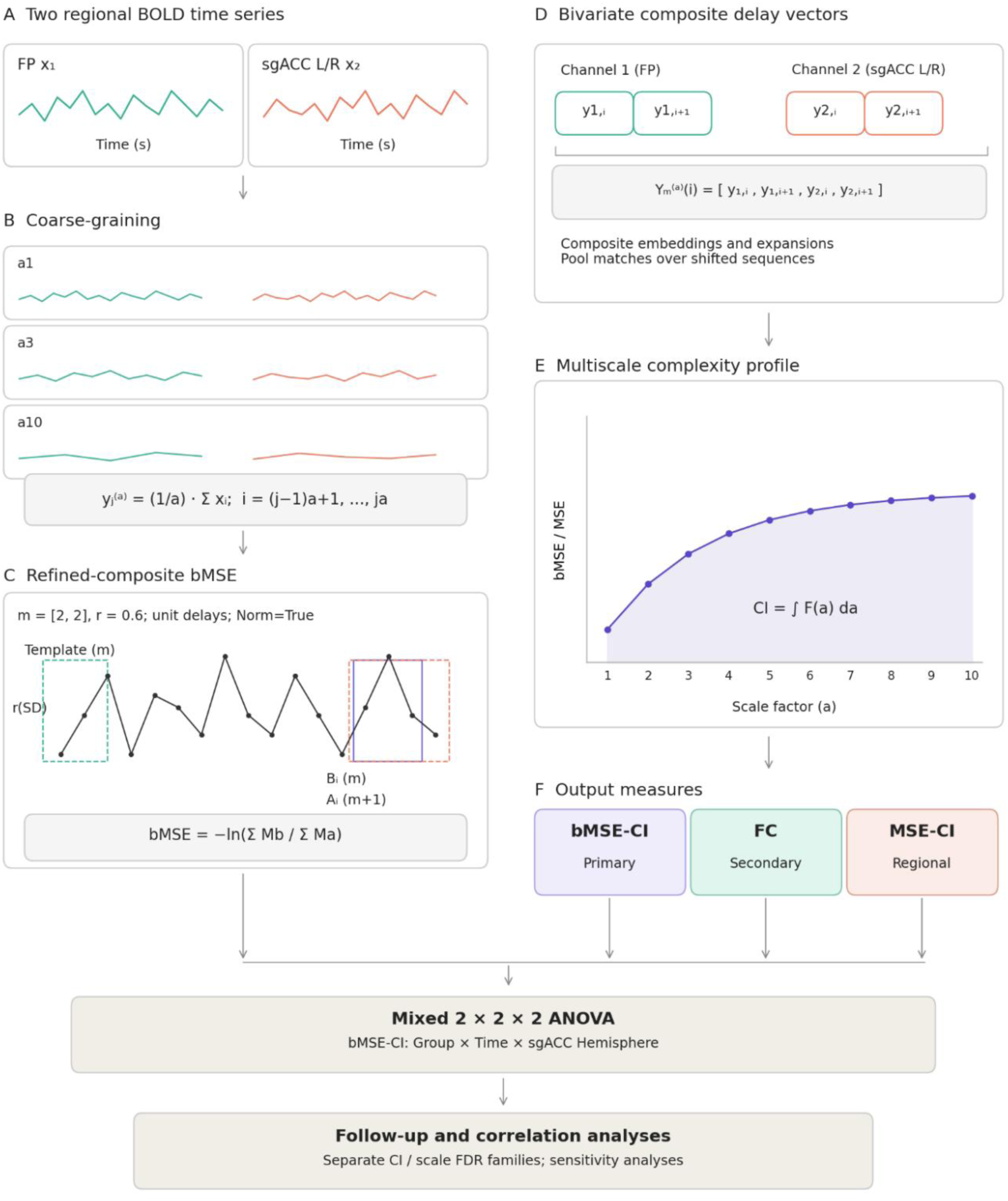
Schematic overview of the FP–sgACC bMSE analysis pipeline. (A) Regional mean BOLD signals from the same bilateral FP mask and either left or right sgACC. (B) Temporal coarse-graining across a1–a10; these are aggregation factors rather than isolated physiological frequency bands. (C–D) Refined-composite bMSE: multivariate embeddings with m = [2, 2], unit delays, r = 0.6 and Norm=True, pooling match probabilities over shifted coarse-grained sequences. The template-matching sketch is illustrative. (E) Entropy profiles and CI obtained by trapezoidal integration across all ten scales. (F) bMSE-CI, FC and voxelwise regional MSE-CI readouts. The primary bMSE-CI test is Group × Time × sgACC Hemisphere; follow-up comparisons, sensitivity models and Spearman correlations use the correction families defined in the methods.

Pearson functional connectivity: 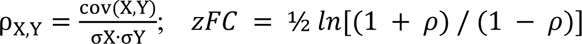 and Y are the FP and respective sgACC regional mean signals; zFC is used in statistical models.

The bMSE complexity index (bMSE-CI) was calculated by trapezoidal integration of the ten scale-specific entropy values with unit spacing across a1–a10. CI summarizes the scale profile; it does not preserve the full profile or quantify inter-scale dependencies. The primary test in this analysis compares FP–left sgACC and FP–right sgACC bMSE-CIs through the Group × Time × Hemisphere interaction. The bilateral sgACC mean-signal bMSE-CI was a secondary readout. Static FC was calculated as the Pearson correlation between denoised FP and each sgACC regional mean signal; statistical models used Fisher-z-transformed FC. Regional univariate MSE was calculated separately at each voxel using non-overlapping coarse-graining, embedding dimension m = 2, unit delay, and tolerance 0.3 times the original, pre-coarse-graining voxel-signal standard deviation (Costa et al., 2002; Costa et al., 2005; Richman & Moor-man, 2000). At each scale, sample entropy was −ln(A/B), where A and B were the counts of distinct unordered matching pairs of length-(m+1) and length-m templates, respectively; counts were not separately normalized by the numbers of possible pairs. Constant signals or zero-match estimates were treated as missing rather than replaced with zero. Regional scale profiles averaged estimable voxel entropies within FP or the respective sgACC mask, and regional MSE-CIs were their trapezoidal integrals. Valid voxel counts were retained for each scale. MSE calculated directly from a regional mean time series was stored separately and is distinct from this voxelwise regional average.

Regional univariate entropy definitions: X_m_(i) = {x_i_, x_i+1_, …, x_i+m−1_}; d[X_m_(i), X_m_(j)] = r; 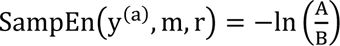. Templates here are formed from the coarse-grained voxel signal; A and B count distinct unordered matching template pairs, with m = 2 and r = 0.3 × SD of the original voxel signal.

Complexity index (unit scale spacing): *CI* = *Σ*_a=1_^9^ [*E*(*a*) + *E*(*a* + 1)] / 2. E(a) denotes bMSE at scale a, or the regional average of estimable voxelwise MSE values for regional MSE-CI.

The left/right analysis contained 156 observations (39 participants × two timepoints × two sgACC sides), with 234 profiles when the secondary bilateral readout was included. Left/right scale-specific bMSE values ranged from 0.392 to 1.449, and bMSE-CIs from 6.495 to 9.460. All exported left/right bMSE profiles, CIs, FC values and regional MSE summary profiles were finite. Individual voxelwise MSE estimates could be nonestimable; those values were excluded from the scale-specific regional average and their valid counts retained.

### Statistical analysis

Analyses used Python 3 with NumPy, SciPy, pandas, statsmodels and Pingouin. The primary test of this analysis was the Group × Time × sgACC Hemisphere interaction for bMSE-CI in a mixed 2 × 2 × 2 ANOVA. Group (patients, controls) was between participants; Time (baseline, follow-up) and Hemisphere (supplied left, supplied right sgACC) were within participants. The same bilateral FP signal was used for both hemispheres. Orthonormal within-participant contrast strata and equal-group Type-III hypotheses were used. With two levels per within-participant factor, sphericity correction was unnecessary. The single primary three-way test used α = 0.05 without multiplicity correction; other ANOVA effects were secondary descriptive tests. The primary interaction was independently checked using a pooled independent-groups t-test of each participant’s contrast [(follow-up − baseline)right − (follow-up − baseline)left], and a mixed ANOVA on right-minus-left CI at each time. Contrast coefficients express controls minus patients. Sensitivity analyses regressed the participant-level contrast on group, age and sex, and additionally ΔFD. Full-factorial random-intercept-only linear mixed-effects models, estimated by restricted maximum likelihood, included Group × Time × Hemisphere, age and sex, with and without session-specific FD. These models impose a more restrictive within-participant covariance structure than the contrast-stratum ANOVA and were reported as model-specification sensitivities. Handedness sensitivities separately excluded documented left-handers or retained only documented right-handers; missing handedness was not imputed.

Participant-level interaction contrast: *D*_i_ = (*CI*_i_, *FU*, *R* − *CI*_i_, *BL*, *R*) − (*CI*_i_, *FU*, *L* − *CI*_i_, *BL*, *L*); *βGroup* = *mean*(*D_controls_*) − *mean*(*D_patients_*). BL and FU denote baseline and follow-up. Comparing D between groups tests the Group × Time × Hemisphere interaction.

Exploratory three-way interactions were tested separately at a1–a10, with Benjamini–Hochberg false discovery rate (FDR) correction across all ten interaction p-values. Secondary Group × Time random-intercept models were fitted separately for FP–left sgACC, FP–right sgACC and FP–bilateral sgACC bMSE-CI, scale-specific bMSE, Fisher-z FC, and voxelwise regional MSE-CIs, adjusting for age and sex and, in separate sensitivity models, session-specific FD. Secondary bMSE-CI interactions formed a three-readout FDR family; scale-specific Group × Time interactions formed ten-test families within each readout. FD-adjusted models used separate correction families. Boundary fits with an invalid joint Hessian were retained using a documented zero-random-variance generalized least-squares limit. Simple effects used paired t-tests for follow-up-minus-baseline changes and Welch t-tests for controls-minus-patients comparisons at each time. For each of bMSE-CI, FC-z and sgACC MSE-CI, the eight left/right simple effects formed a separate FDR family; other exported simple-effect p-values were descriptive and uncorrected. Paired t statistics and dz followed the follow-up-minus-baseline convention, with dz equal to the mean paired difference divided by its sample standard deviation. These simple effects do not replace the direct three-way interaction test. Motion was modeled using one FD value per scan, without duplicating hemispheric observations.

Exploratory patient Spearman correlations related MADRS change to baseline bMSE and bMSE change. All change scores were defined as follow-up minus baseline. The four left/right × baseline/change bMSE-CI correlations formed one Benjamini–Hochberg FDR family. For scale-specific correlations, the twenty left/right × a1–a10 tests formed one family for baseline associations and a separate family for change associations. Bilateral sgACC readouts and each handedness sensitivity were corrected in separately identified families: two CI tests and ten scale tests per baseline/change analysis for the bilateral readout, and the same four-CI/twenty-scale structure for each handedness selection. CI correlation confidence intervals used 5,000 paired-participant bootstrap resamples. Correlations stratified by remission status (MADRS < 10 at follow-up) were descriptive exploratory summaries rather than tests of differential prediction (Hawley et al., 2002).

Associations of Fisher-z FC with bMSE, voxelwise regional FP MSE with bMSE, voxelwise regional sgACC MSE with bMSE, and FP MSE with sgACC MSE were examined using Spearman correlations. Baseline analyses included all participants; change-score analyses included patients only. Each relationship, sgACC readout and baseline/change analysis formed a separate eleven-test FDR family comprising the CI and all ten scales. These null-hypothesis tests do not establish independence between measures.

Exploratory test–retest reliability in the 15 untreated controls was assessed using two-way random-effects, absolute-agreement, single-measurement ICC(2,1), separately for each sgACC readout, bMSE-CI and all ten bMSE scales, FC-z, and regional MSE-CIs and scales. The 95% percentile confidence intervals used 5,000 paired-participant bootstrap resamples (seed = 20261015). Negative estimates and confidence limits were retained. Mean retest differences and limits of agreement were also calculated. Voxelwise MSE estimability was summarized using the valid-voxel count at each scale; nonestimable values were not replaced by zero. Control test–retest estimates do not validate individual sensitivity to treatment-related change.

## Results

Patients showed significant clinical improvement following aTMS treatment, with MADRS scores decreasing from 26.3 ± 9.3 at baseline to 13.8 ± 8.5 at follow-up (t(23) = −6.250, p < 0.001). Nine of 24 patients (37.5%) met criteria for remission (MADRS < 10) at follow-up. Clinical characteristics of the present analysis sample are summarized in Table 1; clinical results for the smaller connectivity analysis sample were reported by Bracht et al. (2026).

**Table 1:**
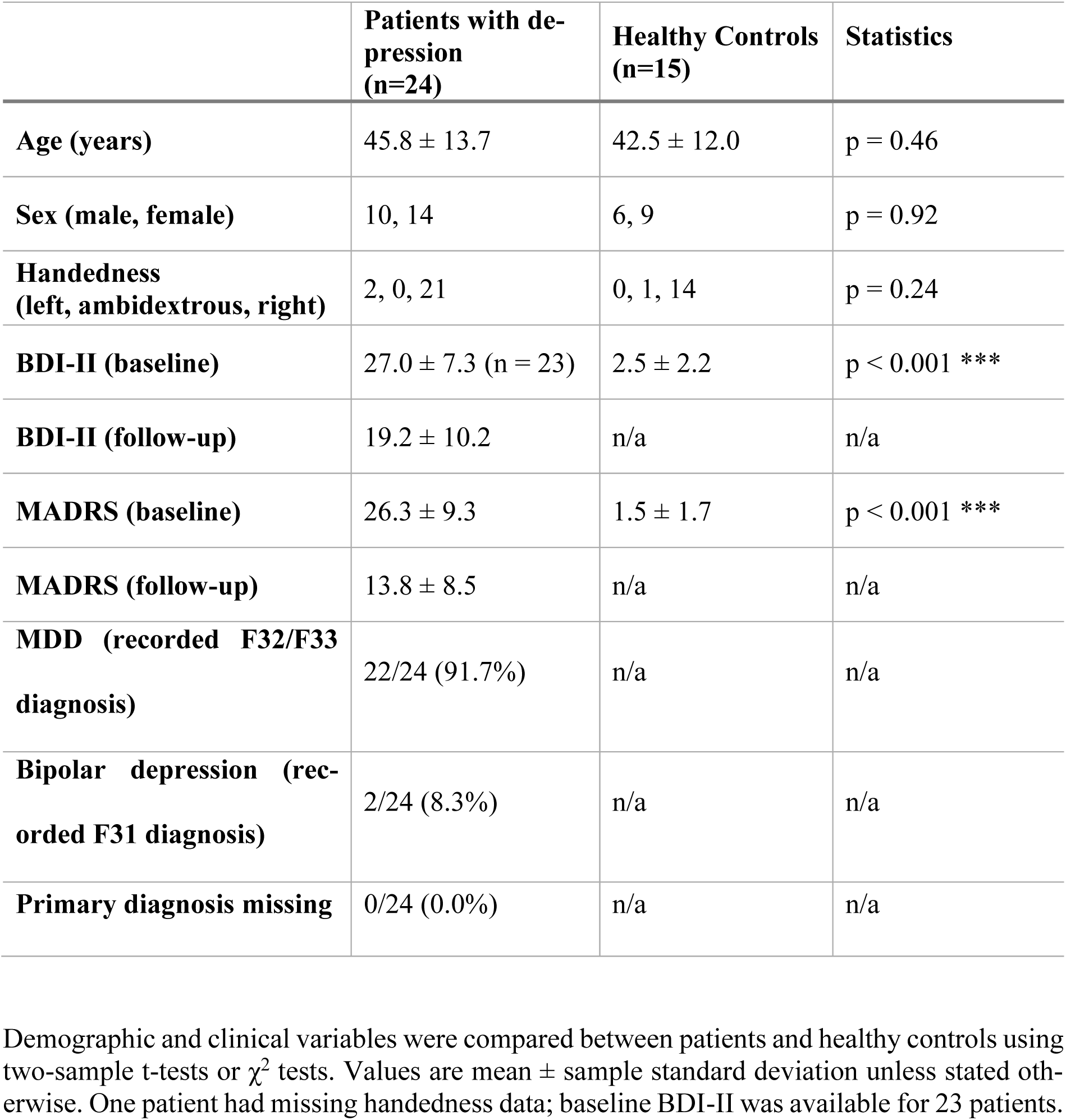
Clinical characteristics of the present analysis sample.

|  | Patients with de-pression<br>(n=24) | Healthy Controls (n=15) | Statistics |
|---|---|---|---|
| **Age (years)** | 45.8 ± 13.7 | 42.5 ± 12.0 | p = 0.46 |
| **Sex (male, female)** | 10, 14 | 6, 9 | p = 0.92 |
| **Handedness<br>(left, ambidextrous, right)** | 2, 0, 21 | 0, 1, 14 | p = 0.24 |
| **BDI-II (baseline)** | 27.0 ± 7.3 (n = 23) | 2.5 ± 2.2 | p < 0.001 *** |
| **BDI-II (follow-up)** | 19.2 ± 10.2 | n/a | n/a |
| **MADRS (baseline)** | 26.3 ± 9.3 | 1.5 ± 1.7 | p < 0.001 *** |
| **MADRS (follow-up)** | 13.8 ± 8.5 | n/a | n/a |
| **MDD (recorded F32/F33 diagnosis)** | 22/24 (91.7%) | n/a | n/a |
| **Bipolar depression (rec-orded F31 diagnosis)** | 2/24 (8.3%) | n/a | n/a |
| **Primary diagnosis missing** | 0/24 (0.0%) | n/a | n/a |

Demographic and clinical variables were compared between patients and healthy controls using two-sample t-tests or χ<sup>2</sup> tests. Values are mean ± sample standard deviation unless stated oth-erwise. One patient had missing handedness data; baseline BDI-II was available for 23 patients.

### FP–sgACC bMSE-CI: Group × Time × Hemisphere

The primary exploratory Group × Time × sgACC Hemisphere interaction for bMSE-CI was significant (F(1,37) = 6.043, p = 0.0188, partial η^2^ = 0.140; Figure 3). The control-minus-patient difference in the right-minus-left change contrast was β = −0.620 (95% CI −1.132 to −0.109). The other six bMSE-CI ANOVA effects did not reach p < 0.05; in particular, the Group × Time effect averaged across hemispheres was not significant (p = 0.207).

**Figure 3:**
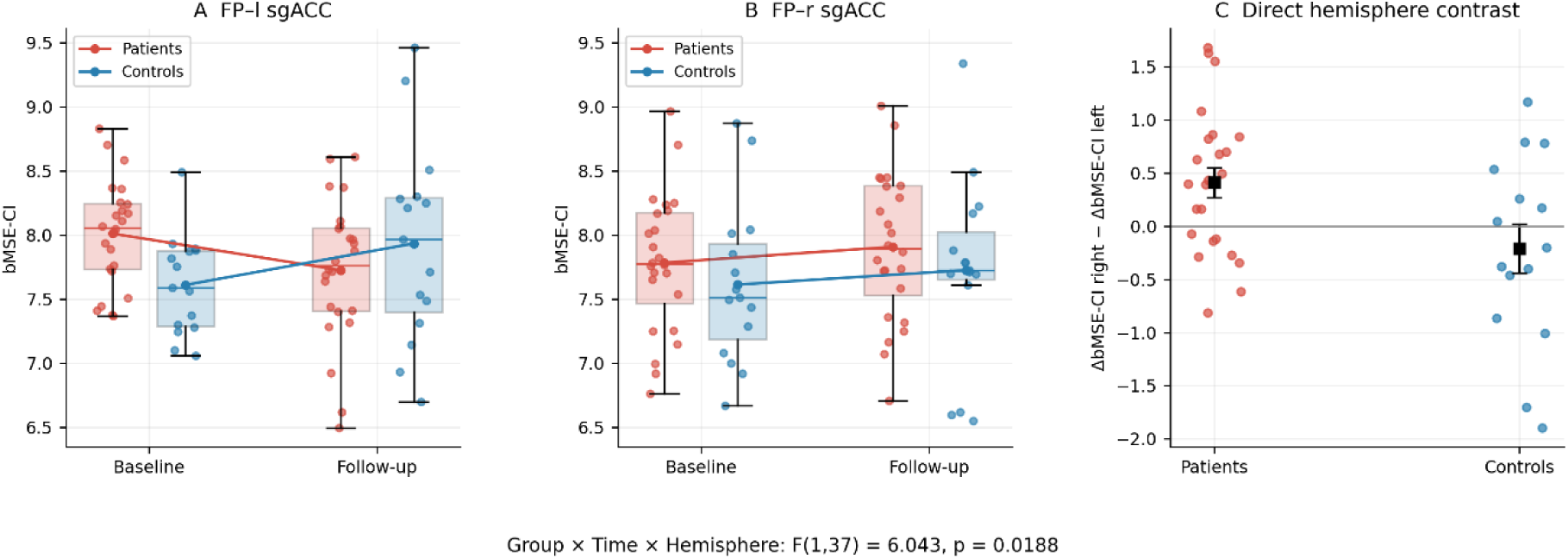
Longitudinal FP–sgACC bMSE-CI and the direct hemispheric change contrast. (A) FP–left sgACC and (B) FP–right sgACC bMSE-CI at baseline and follow-up for patients (red; n = 24) and controls (blue; n = 15). Dots represent individual observations. Boxes show the median and interquartile range, with whiskers to observations within 1.5 interquartile ranges; lines connect group means. A and B share the same y-axis limits. (C) Individual right-minus-left change contrasts, [(follow-up − baseline)right − (follow-up − baseline)left], with group means ± SEM shown as black squares. The Group × Time × Hemisphere interaction was F(1,37) = 6.043, p = 0.0188. The hemispheric comparison uses the identical bilateral FP signal for both sgACC sides.

For FP–left sgACC, baseline-to-follow-up bMSE-CI was patients: 8.010 ± 0.412 to 7.724 ± 0.551; controls: 7.609 ± 0.389 to 7.933 ± 0.786. For FP–right sgACC, baseline-to-follow-up bMSE-CI was patients: 7.783 ± 0.545 to 7.907 ± 0.578; controls: 7.612 ± 0.624 to 7.724 ± 0.735. The three-way participant-contrast result remained significant after age/sex adjustment (β = −0.652, 95% CI −1.170 to −0.134, p = 0.0152) and additional ΔFD adjustment (β = −0.637, 95% CI −1.182 to −0.093, p = 0.0231). The full-factorial random-intercept-only mixed model gave p = 0.0718, and p = 0.0720 with FD; the inference therefore depended on covariance-model specification.

### Scale-specific three-way interactions

Only a2 survived FDR correction across the ten scale-specific Group × Time × Hemisphere tests (F(1,37) = 9.229, p = 0.004351, q = 0.0435); the remaining scales had q ≥ 0.134 (Figure 4). This scale factor denotes temporal aggregation rather than an isolated physiological frequency band. The secondary age/sex-adjusted left-readout Group × Time CI interaction was β = 0.609 (95% CI 0.160 to 1.058; p = 0.0079; q = 0.0237). The secondary age/sex-adjusted right-readout Group × Time CI interaction was β = −0.011 (95% CI −0.553 to 0.530; p = 0.9674; q = 0.9674). The secondary age/sex-adjusted bilateral-readout Group × Time CI interaction was β = 0.510 (95% CI −0.063 to 1.083; p = 0.0810; q = 0.1215). Separate readout-specific tests do not themselves establish a hemispheric difference. No within-group CI change survived the eight-test left/right simple-effects FDR correction; the left-pair baseline control-minus-patient comparison did (mean difference = −0.400, q = 0.0368). The bilateral mean-signal readout is secondary and is not equivalent to the three-way hemispheric test. Unadjusted participant-con-trast handedness sensitivities remained significant when excluding documented left-handers (n = 37, p = 0.0264) or retaining documented right-handers only (n = 35, p = 0.0126).

**Figure 4:**
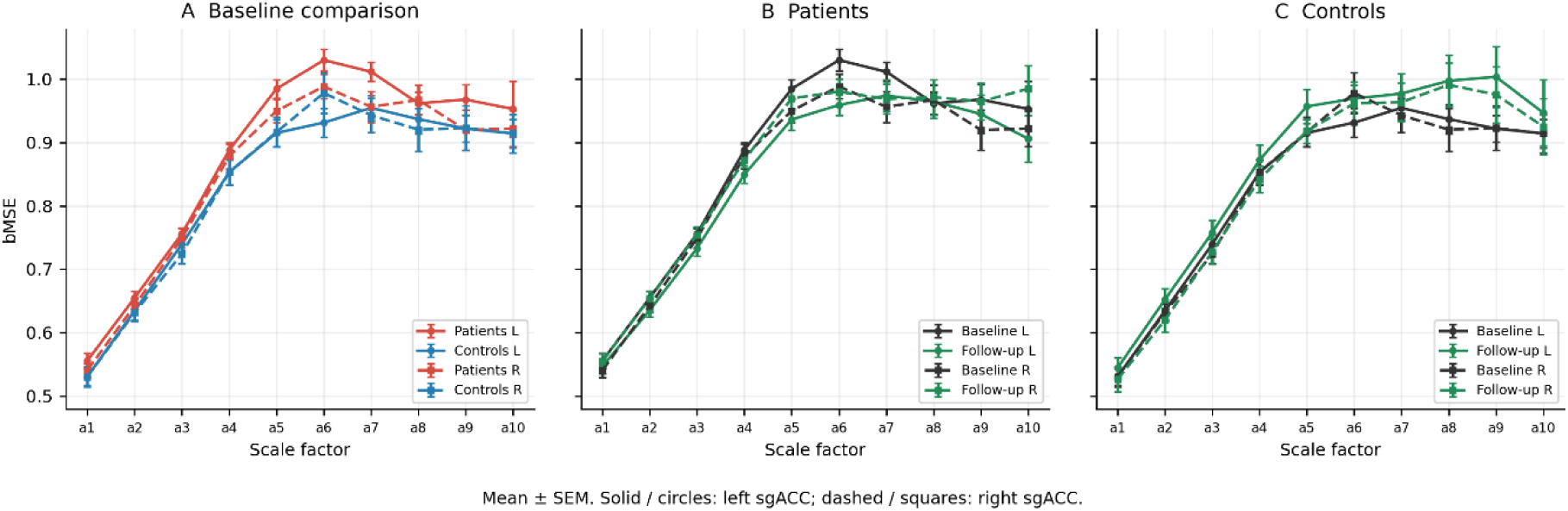
Scale-specific FP–sgACC bMSE profiles. Three panels in one row show (A) the baseline group comparison, (B) patients at baseline and follow-up, and (C) controls at baseline and follow-up. Solid lines with circles denote left sgACC; dashed lines with squares denote right sgACC. In A, patients are red and controls blue; in B and C, baseline is black and follow-up green. All panels use the same axis limits; points and error bars show means ± SEM. No within-group significance markers are used as evidence for a hemispheric interaction. Across the ten Group × Time × Hemisphere tests, a2 had FDR q = 0.0435. Scale factors a1–a10 describe temporal aggregation.

### Functional connectivity (secondary outcome)

No statistically significant age/sex-adjusted Group × Time interaction was detected for Fisher-z FP–sgACC FC: left, β = 0.003, 95% CI −0.139 to 0.145, p = 0.969; right, β = 0.047, 95% CI −0.086 to 0.181, p = 0.487. These are secondary within-readout models rather than a direct FC hemispheric interaction test.

### Voxelwise regional MSE (secondary outcome)

The voxelwise regional FP MSE-CI showed no statistically significant Group × Time interaction (β = 0.073, 95% CI −0.014 to 0.161, p = 0.100). The FP signal and regional measure were identical for the left and right sgACC pairs.

No statistically significant Group × Time interaction was detected for voxelwise regional sgACC MSE-CI: left, β = 0.052, 95% CI −0.048 to 0.153, p = 0.308; right, β = 0.062, 95% CI − 0.033 to 0.156, p = 0.200. The difference between significant bMSE and nonsignificant FC/MSE tests does not establish superiority of bMSE.

### Exploratory clinical associations

Patient MADRS change was not significantly associated with bMSE-CI change for either sgACC pair (left: ρ = −0.113, 95% bootstrap CI −0.496 to 0.314, p = 0.600, q = 0.937; right: ρ = 0.029, 95% bootstrap CI −0.448 to 0.495, p = 0.892, q = 0.937). The four left/right base-line/change CI tests formed one FDR family (Figure 5).

**Figure 5:**
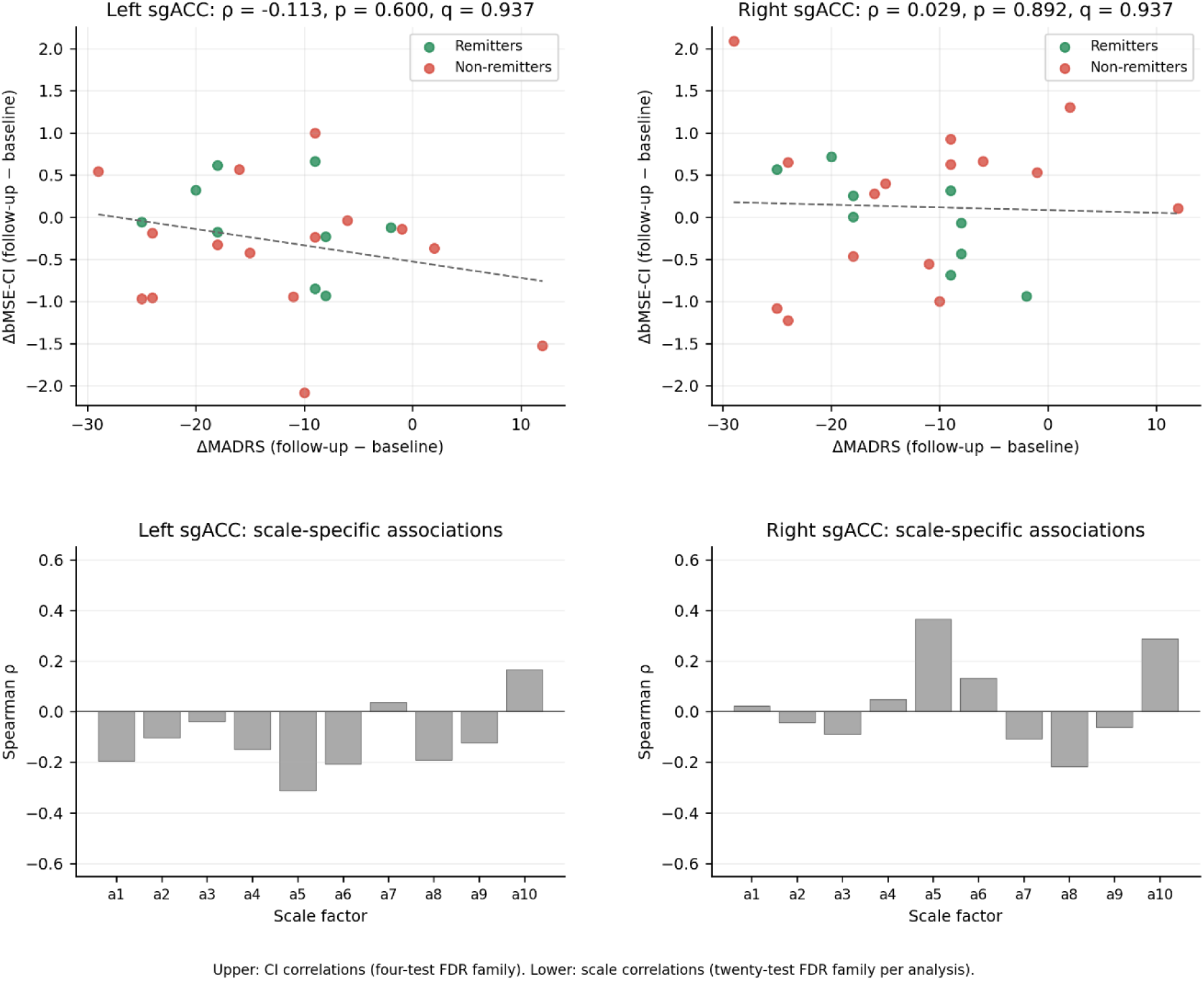
Associations between MADRS change and FP–sgACC bMSE change in patients. Left and right columns show the left and right sgACC pairs, respectively (n = 24 patients each). Upper panels show ΔMADRS versus ΔbMSE-CI, with remitters (follow-up MADRS < 10) in green and non-remitters in red. Dashed lines are descriptive least-squares fits; annotations give Spearman ρ, raw p and FDR q. The four CI tests across left/right and baseline/change form one FDR family. Lower panels show scale-specific ΔMADRS–ΔbMSE correlations at a1–a10; red bars denote q < 0.05, otherwise bars are gray. The twenty left/right scale-change correlations form one FDR family. All changes are follow-up minus baseline; negative ΔMADRS indicates symptom improvement.

No scale-specific clinical association survived FDR correction in either the baseline or change-score family, each comprising twenty left/right × scale tests. Remission-stratified correlations were descriptive and did not establish differential predictive performance.

Patient MADRS change was not significantly associated with baseline bMSE-CI for either sgACC pair (left: ρ = 0.076, 95% bootstrap CI −0.250 to 0.379, p = 0.724, q = 0.937; right: ρ = −0.017, 95% bootstrap CI −0.440 to 0.401, p = 0.937, q = 0.937). The four left/right base-line/change CI tests formed one FDR family (Figure 6).

**Figure 6:**
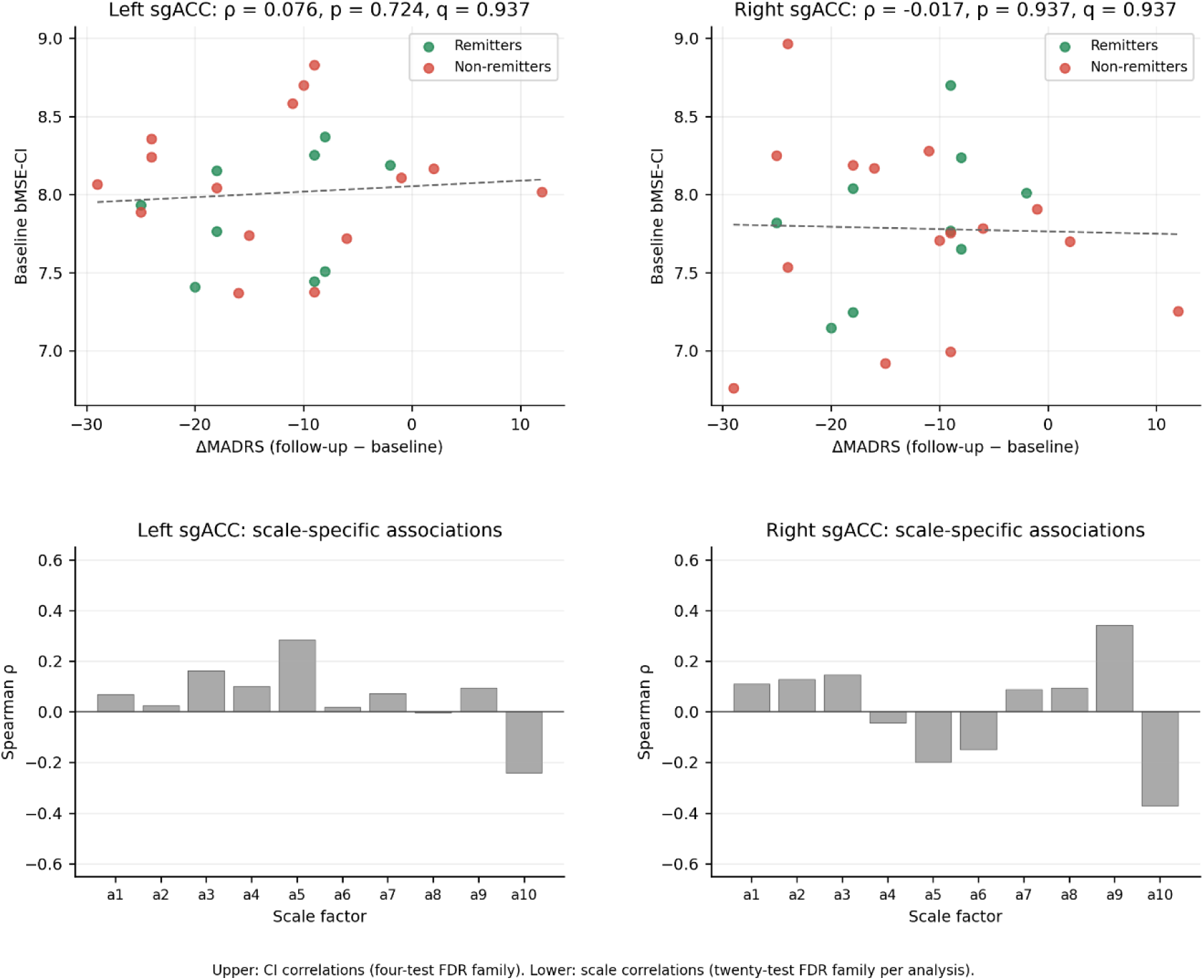
Associations between MADRS change and baseline FP–sgACC bMSE in patients. Left and right columns show the left and right sgACC pairs, respectively (n = 24 patients each). Upper panels show ΔMADRS versus baseline bMSE-CI, colored by remission status (green: remitters; red: non-remitters). Dashed lines are descriptive least-squares fits; annotations give Spearman ρ, raw p and FDR q from the same four-test CI family used in Figure 5. Lower panels show scale-specific correlations between ΔMADRS and baseline bMSE at a1–a10; red bars denote q < 0.05, otherwise bars are gray. The twenty left/right baseline-scale correlations form a separate FDR family from change-score correlations. ΔMADRS is follow-up minus baseline.

### Associations between functional connectivity and joint complexity

For baseline FP–right sgACC, FC-z correlated with bMSE at a1 (ρ = −0.582, p = 0.000102, q = 0.0011; Figure 7). No other left/right baseline FC–bMSE association survived its complete eleven-test CI/scale FDR family.

**Figure 7:**
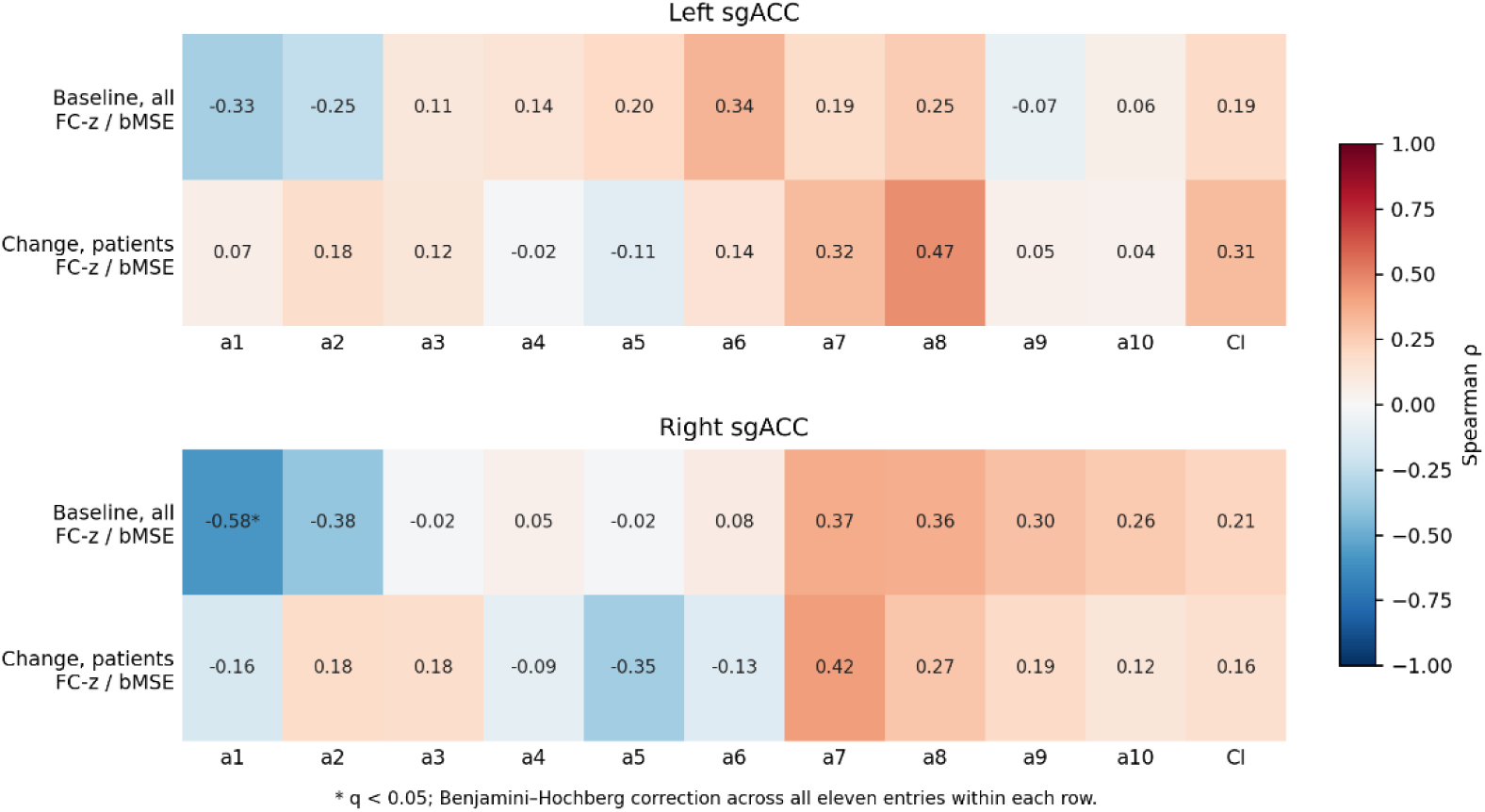
Correlations between FP–sgACC functional connectivity and bMSE. Heatmaps are displayed separately for left sgACC (upper) and right sgACC (lower). Each heatmap shows baseline associations in all 39 participants and change-score associations in 24 patients. Columns show a1–a10 and CI; cells contain Spearman coefficients between FC-z and the corresponding bMSE measure. Colors range from negative (blue) to positive (red), with identical limits of −1 to +1. Asterisks indicate q < 0.05 after Benjamini–Hochberg correction across all eleven entries within each row. ΔFC-z and ΔbMSE are follow-up minus baseline.

No change-score FC–bMSE association survived FDR correction in patients for either sgACC side. These null tests do not establish independence between FC and joint temporal complexity.

### Associations between joint and regional complexity

At baseline across all participants, no tested relationship between voxelwise regional MSE and bMSE, or between FP and sgACC regional MSE, survived the eleven-test FDR correction applied separately to each relationship and sgACC side (Figure 8). This does not establish statistical independence between the measures.

**Figure 8:**
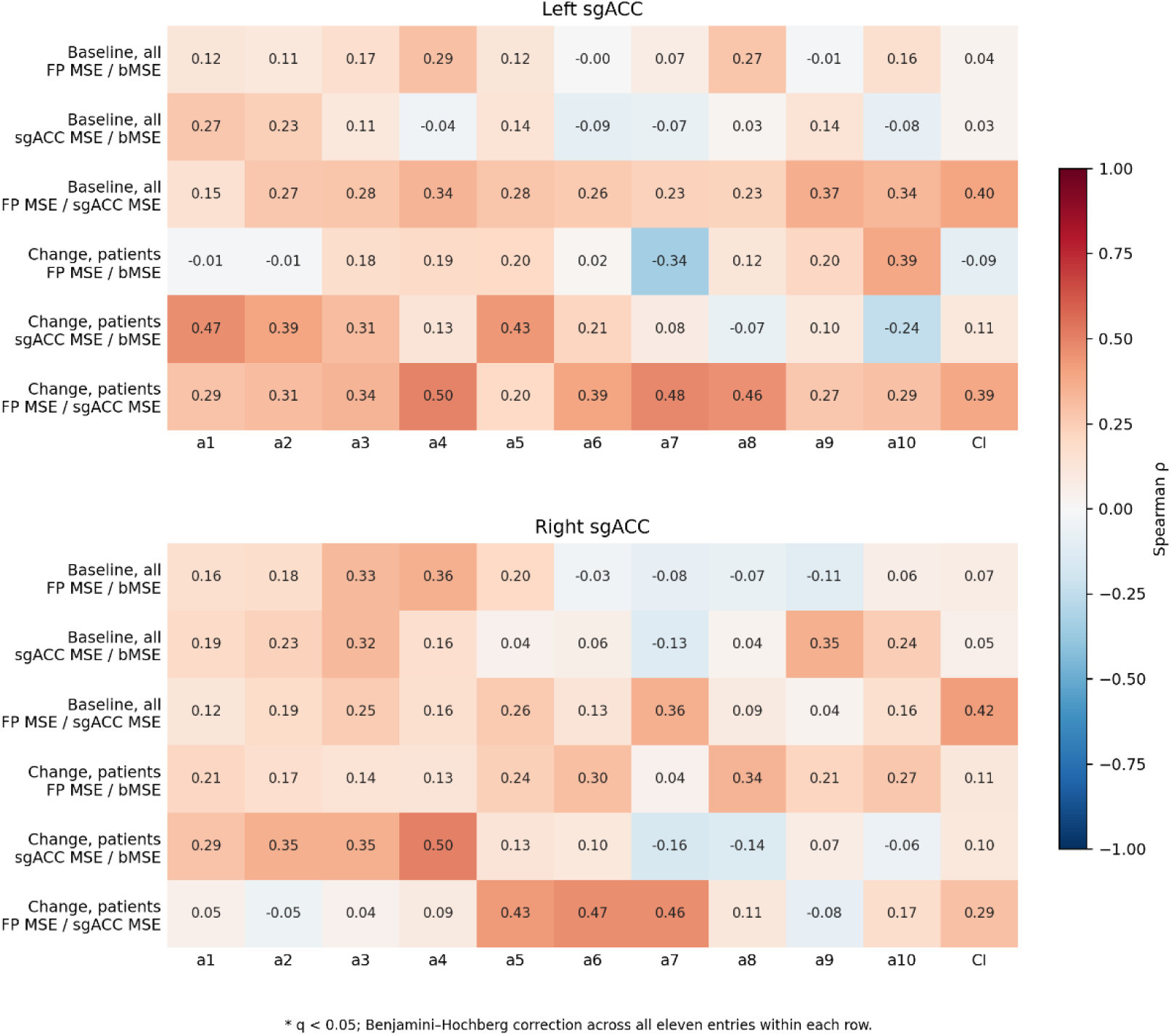
Correlations between FP–sgACC bMSE and voxelwise regional MSE. Heatmaps are displayed separately for left sgACC (upper) and right sgACC (lower). Within each heatmap, the first three rows show baseline correlations in all 39 participants and the last three change-score correlations in 24 patients. Relationships are FP regional MSE versus bMSE, sgACC regional MSE versus bMSE, and FP regional MSE versus sgACC regional MSE. Regional MSE profiles average estimable voxel entropies. Columns show a1–a10 and CI; cells contain Spearman coefficients. Identical color limits of −1 to +1 are used. Asterisks indicate q < 0.05 after Benjamini–Hochberg correction across all eleven entries within each row. All change scores are follow-up minus baseline.

For change scores in patients, no tested relationship between voxelwise regional MSE and bMSE, or between FP and sgACC regional MSE, survived the eleven-test FDR correction applied separately to each relationship and sgACC side (Figure 8). This does not establish statistical independence between the measures.

In the 15 untreated controls, FP–left sgACC bMSE-CI ICC(2,1) = 0.422 (95% bootstrap CI 0.115 to 0.653); mean retest difference = 0.323 ± 0.642, with limits of agreement −0.935 to 1.582; FP–right sgACC bMSE-CI ICC(2,1) = −0.141 (95% bootstrap CI −0.451 to 0.122); mean retest difference = 0.112 ± 1.026, with limits of agreement −1.898 to 2.123. Negative estimates were retained. These uncertain estimates do not validate individual treatment-change measurement.

## Discussion

This study identified an exploratory group-dependent hemispheric difference in longitudinal FP–sgACC joint temporal complexity during aTMS. Mean bMSE-CI decreased for the left sgACC pair and increased for the right pair in patients, whereas controls showed increases on both sides. Because the same bilateral FP signal was used in both pairs, the hemispheric comparison concerns the sgACC component. The findings describe longitudinal differences between patients and untreated controls; the observational design does not establish that aTMS caused these differences.

### Interpretation and robustness of hemispheric differences

The direct three-way interaction supports a difference between groups in the hemispheric pattern of change, but its statistical significance depended on model specification. The participant-level contrast remained significant after adjustment for age, sex, and motion change, whereas random-intercept-only mixed models did not reach significance (p ≈ 0.072). These approaches impose different within-participant covariance assumptions, so the results provide qualified evidence for the hemispheric effect. The left-pair baseline group difference and the absence of FDR-significant within-group CI changes also caution against interpreting the interaction as a demonstrated treatment-induced reduction in patients. Replication should assess the longitudinal contrast while accounting for baseline variability.

### Scale-specific hemispheric differences

The a2 interaction survived FDR correction across all ten scales and identifies a difference at a fine temporal aggregation scale. With TR = 1 s, a2 represents a 2-s aggregation window applied to BOLD signals already filtered to 0.008–0.09 Hz; it does not identify an isolated physiological frequency band or establish high-frequency neural activity, neural criticality, or excitation–in-hibition balance. The integrated CI and scale-specific analyses summarize different aspects of the entropy profile, and the a2 result does not imply that this scale alone accounts for the CI interaction.

### Associations between functional connectivity and complexity

The secondary FC and regional MSE analyses detected no Group × Time interactions, while one baseline association between right-pair FC and a1 bMSE survived FDR correction. These results help characterize the measures but do not establish superior sensitivity of bMSE or independence from FC and regional MSE. In particular, the secondary readout-specific tests address different hypotheses from the primary three-way interaction. FC summarizes linear co-variation, whereas bMSE evaluates the joint temporal complexity of embedded regional signals (McDonough & Nashiro, 2014; Wang et al., 2018). Further comparisons are needed to determine whether bMSE adds reproducible information about longitudinal neural change (de Moura Targino et al., 2025). In a smaller analysis sample from the same observational cohort, Bracht et al. (2026) reported reduced sgACC connectivity with bilateral precunei and the left frontal pole following aTMS. That seed-based voxelwise analysis and the present FP–sgACC analyses differ in sample composition, spatial scope, and outcome measures; they therefore do not constitute independent replication or directly equivalent tests.

The present study used bMSE rather than multiscale cross-sample entropy, which compares templates from one signal against patterns in another to quantify inter-signal asynchrony (Jamin & Humeau-Heurtier, 2020). bMSE instead embeds both channels in composite delay vectors, producing a joint-complexity estimate influenced by within-channel dynamics and cross-channel dependencies (Ahmed & Mandic, 2011; Ahmed & Mandic, 2012). It does not isolate directed coupling or information exchange.

### Circuit-level considerations and limitations

Several limitations constrain interpretation. The modest sample size limits precision and generalizability, and the hemispheric analysis was not prospectively prespecified. Test–retest reliability in the 15 controls was uncertain: bMSE-CI ICC estimates were 0.422 for the left pair and −0.141 for the right pair, with broad confidence intervals. These estimates limit confidence in individual change measurement and do not establish suitability for treatment monitoring. No clinical bMSE association survived the specified FDR corrections. The absence of a sham-treated patient group leaves habituation, time effects, and nonspecific treatment effects as alternative explanations. Medication exposure and changes, diagnostic heterogeneity, and residual head motion may also contribute. The unequal spatial extent of FP and sgACC masks may affect signal-to-noise ratio and entropy estimates, while entropy remains sensitive to spectral composition despite the refined-composite estimator. Acquisition at 7 Tesla offers potential signal-to-noise advantages (Pohmann et al., 2016), but susceptibility-related distortion and signal dropout in the sgACC remain possible sources of bias.

In conclusion, bMSE identified an exploratory hemisphere-dependent longitudinal difference in FP–sgACC joint temporal complexity between patients undergoing aTMS and untreated healthy controls. The integrated CI interaction and the FDR-corrected a2 interaction motivate further evaluation of bMSE as a candidate measure of longitudinal neural change in depression. Model-dependent inference, uncertain reliability, and the absence of corrected clinical associations make independent replication and prospective validation necessary before considering applications in treatment monitoring or personalized intervention.

## Data and Code Availability

All data produced in the present study are available upon reasonable request to the authors. The MRIEntropyMetrics software used in this study has been published under the MIT License and is publicly available at https://github.com/brain-complexity/MRIEntropyMetrics.

## Author Contributions

N.D. – Conceptualization, Methodology, Software, Formal Analysis, Investigation, Data Curation, Project Administration, Writing (Orig + Rev), Visualization

M.G. – Resources, Methodology, Writing (Rev)

K.J. – Resources, Methodology, Software, Writing (Rev)

P.B. – Investigation, Data Curation, Project Administration, Writing (Rev)

S.D. – Investigation, Data Curation, Project Administration, Writing (Rev)

G.F. – Resources, Writing (Rev)

K.A. – Resources, Writing (Rev)

A.F. – Resources, Methodology, Writing (Rev)

R.W. – Resources, Writing (Rev)

L.M.S. – Resources, Writing (Rev)

S.W. – Conceptualization, Investigation, Writing (Rev), Funding Acquisition

T.B. – Conceptualization, Investigation, Project Administration, Writing (Orig + Rev), Funding Acquisition

## Data Availability

All data produced in the present study are available upon reasonable request to the authors.

## Acknowledgements

AI-assisted language editing was performed using Perplexity AI (Perplexity AI, Inc., San Francisco, CA, USA). Codex (OpenAI) was also used for language editing, manuscript consistency checks, and reference checks. The authors retain responsibility for the scientific content and the final manuscript.

---

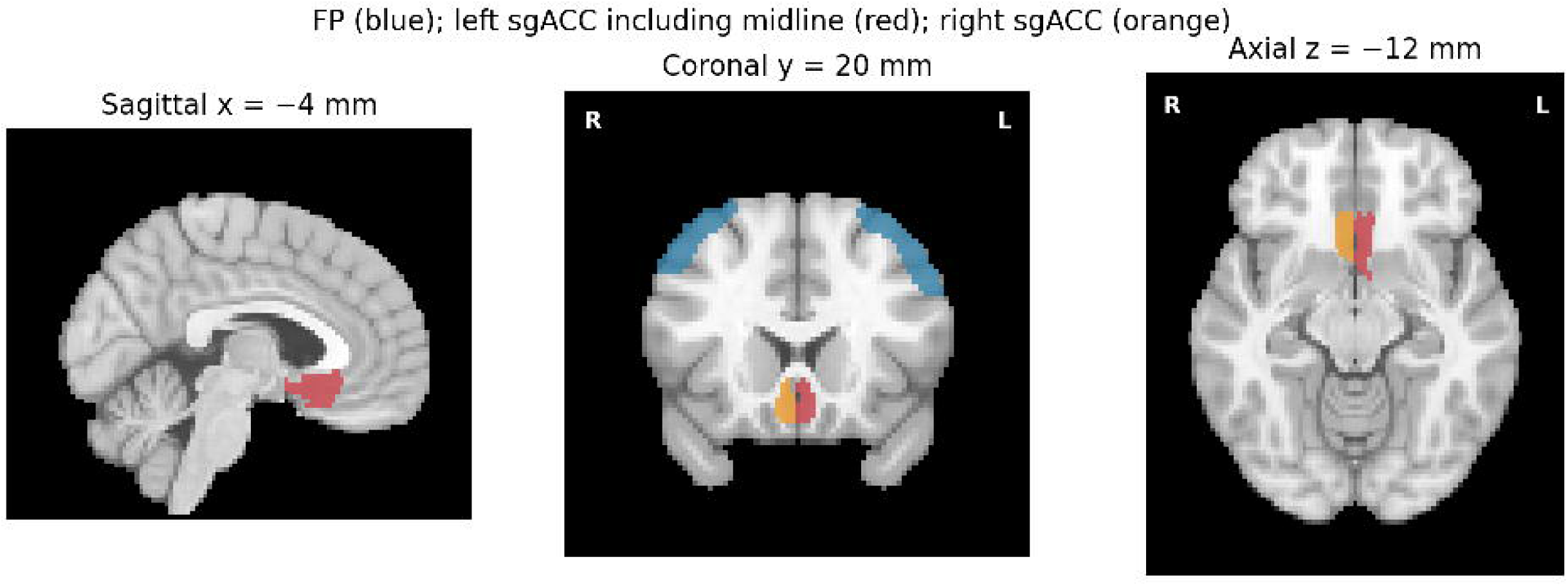

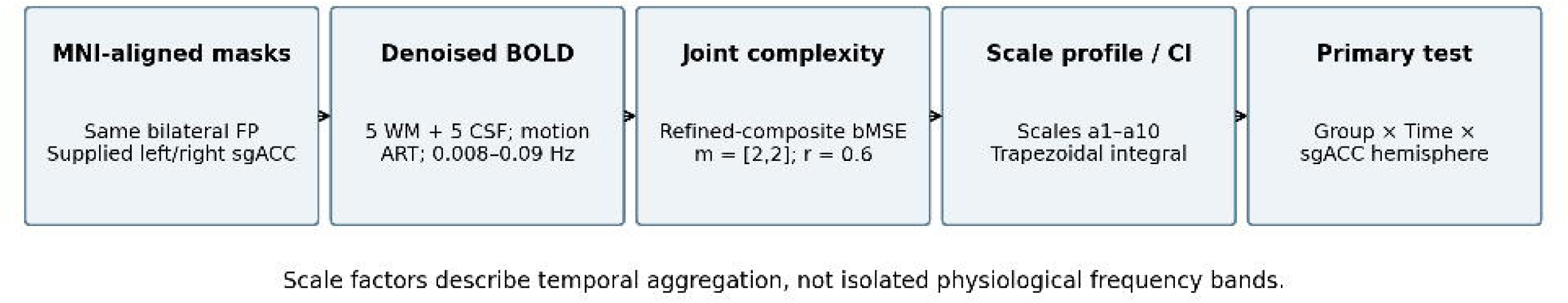

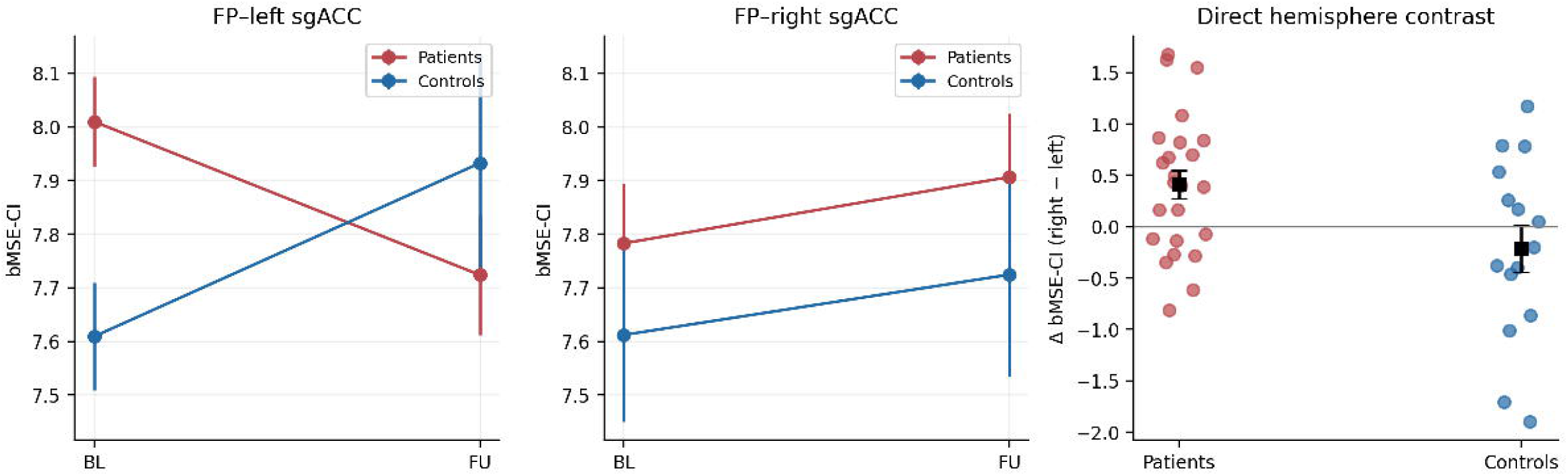

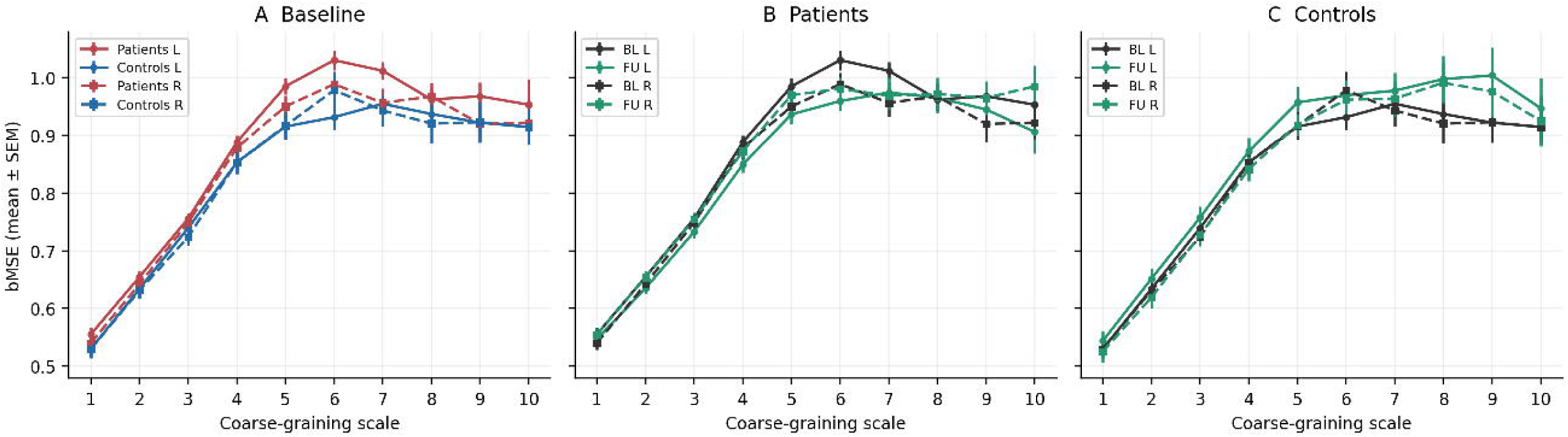

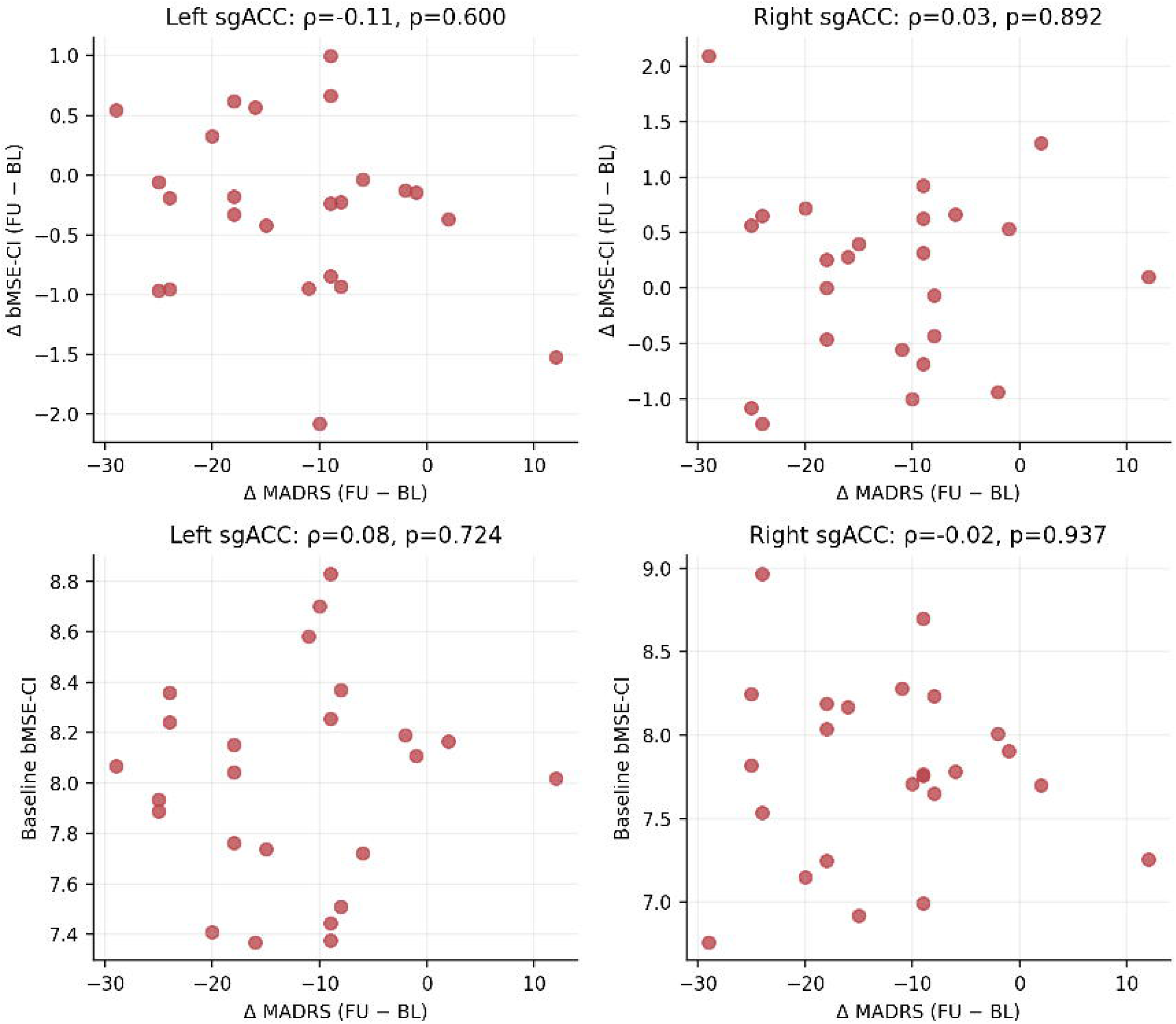

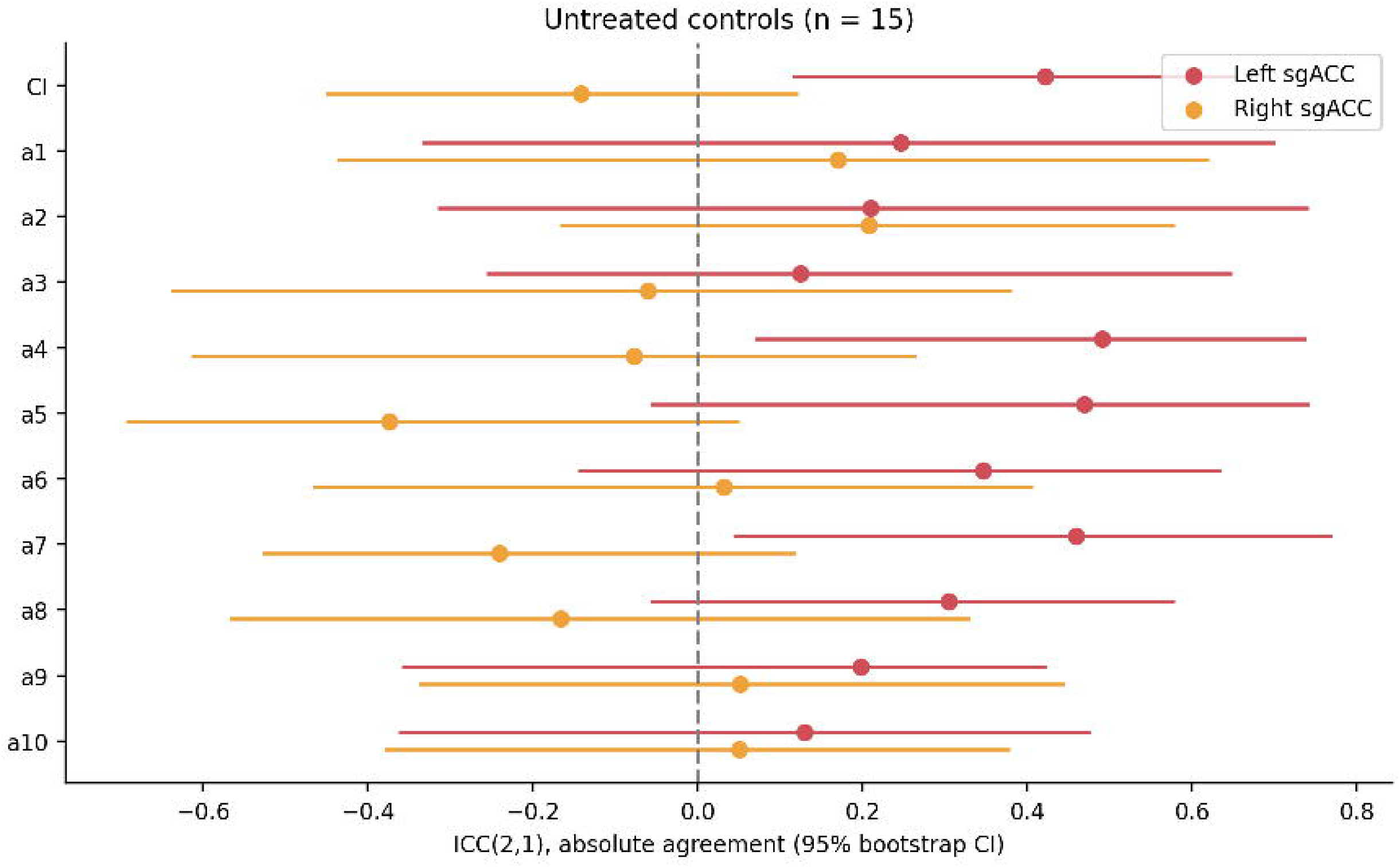

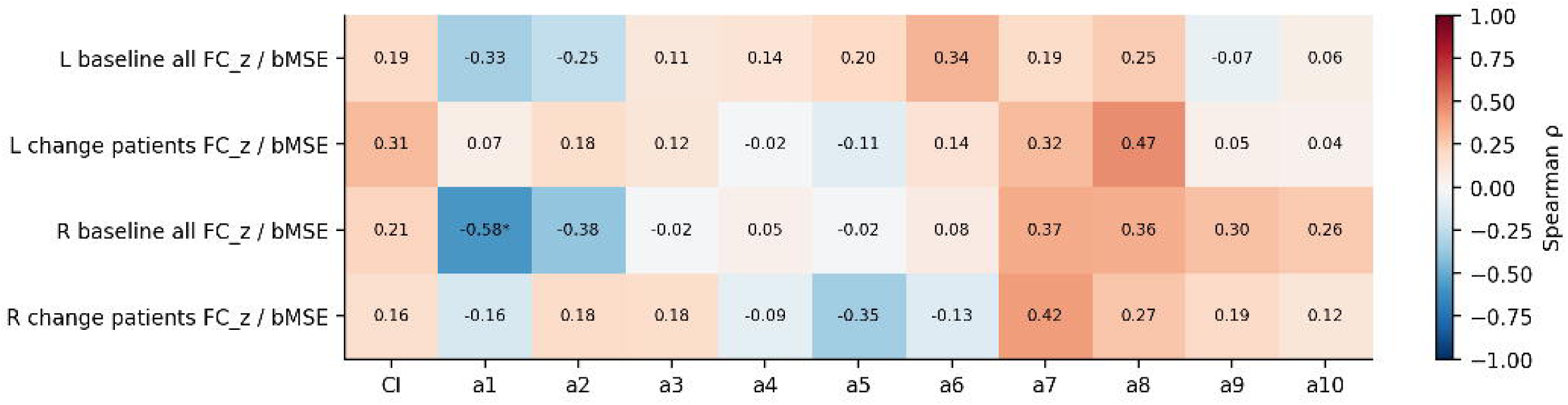

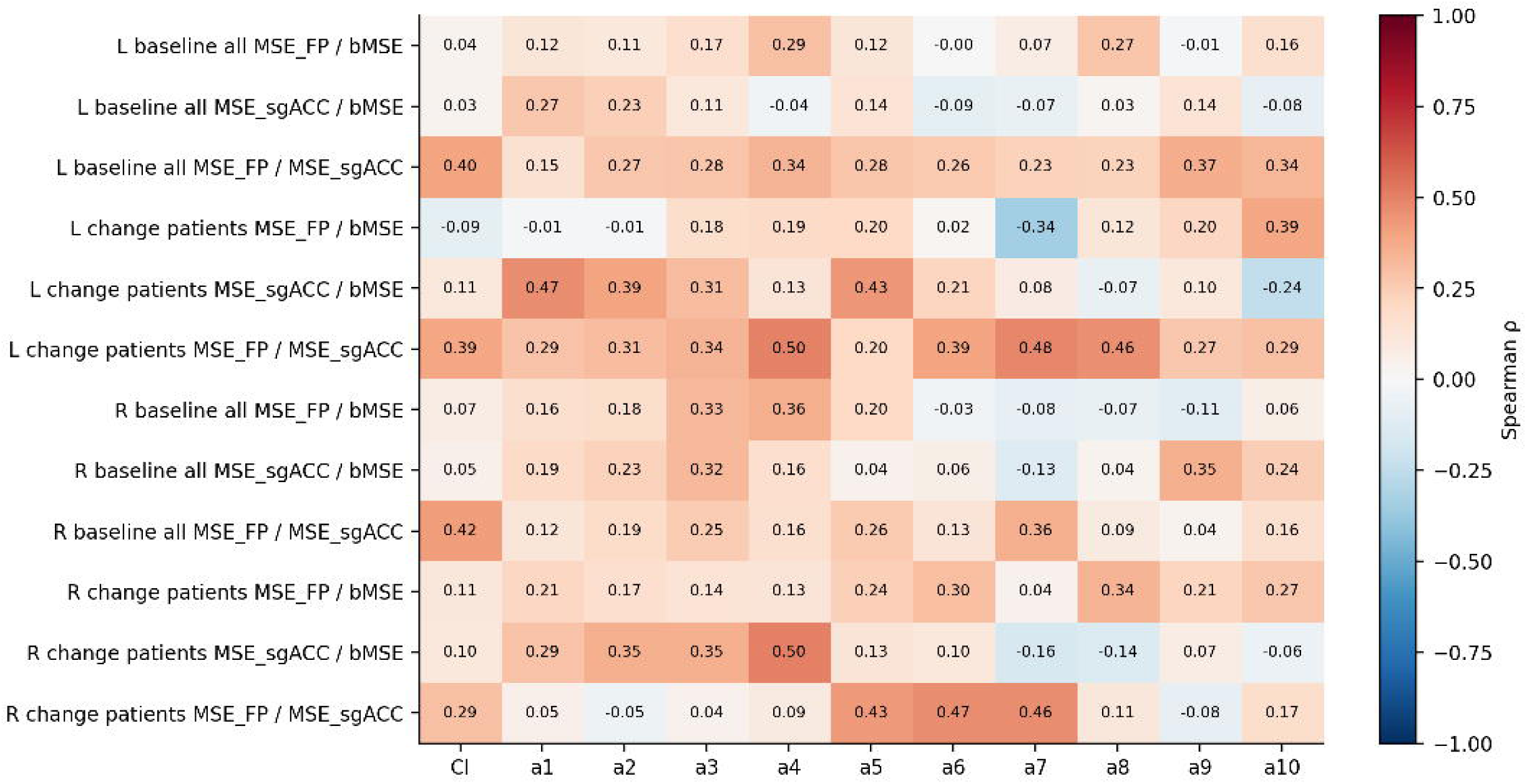

